# Physician Decision-Making in Acute Ischemic Stroke with and without a Guideline-Encoded Clinical Decision Support System

**DOI:** 10.64898/2026.10.26.26366524

**Authors:** Mohamed F. Doheim, Raul G. Nogueira

## Abstract

**Background:** Applying extensive, frequently revised guidelines at the point of care is difficult. We evaluated whether acute ischemic stroke decisions made with LUCID, a clinical decision support system encoding the 2026 American Heart Association/American Stroke Association guideline, were more often guideline-concordant than decisions made without it.

**Methods:** In this pilot vignette study, clinicians completed 8 stroke vignettes (24 questions) alone (Arm A) or with access to LUCID (Arm B). Questions bundling recommendations of different classes were split into nodes (29 decision points). The primary outcome, full guideline concordance, was compared between decision points made with and without LUCID using cluster-bootstrap risk differences (RDs). Decision confidence and workload (NASA Task Load Index; missing scores multiply imputed) were compared between arms.

**Results:** Fifteen clinicians (Arm A, 5; Arm B, 10) contributed 351 decision points: 151 with LUCID and 200 without. Full concordance was 99.3% (150/151) with LUCID versus 71.0% (142/200) without (RD, 28.3 [95% CI, 21.7–34.9] percentage points; P<0.001), and at least partial concordance was 100% versus 93.5% (RD, 6.5 [2.3–10.2]; P=0.001). Concordance with versus without LUCID was 100% versus 67.9% for Class 1, 100% versus 79.3% for Class 3: Harm, and 100% versus 74.3% for Class 3: No Benefit recommendations. Commission errors occurred at 0 of 66 versus 10 of 64 Class 3 decision points (P<0.001). Decision confidence was numerically higher in Arm B than in Arm A (median, 5 versus 4; P=0.24). Workload was lower in Arm B for mental demand (difference, −2.9 points; P=0.045), with borderline reductions in temporal demand (P=0.090) and composite workload (P=0.084).

**Conclusions:** In this pilot, decisions made with a guideline-encoded decision support system were almost uniformly guideline-concordant, including for recommendations against harmful interventions, and workload tended to be lower with system access. Confirmation in a larger study with blinded outcome assessment is warranted.

## INTRODUCTION

Stroke remains a leading cause of death and long-term disability in the United States and worldwide.^1–3^ The 2026 American Heart Association/American Stroke Association (AHA/ASA) guideline for the early management of acute ischemic stroke revised recommendations across the entire care pathway, from prehospital triage to in-hospital complications, incorporating recent trials of basilar artery thrombectomy, medium- and distal-vessel occlusion, blood pressure targets after thrombectomy, and antithrombotic timing.^4^ Each recommendation is graded by Class of Recommendation (COR) and Level of Evidence (LOE): Class 1 recommendations identify interventions that should be performed, and Class 3 recommendations identify interventions that provide no benefit or cause harm.^5^

Applying an evolving and highly granular guideline consistently at the point of care is difficult. National quality programs have improved adherence to core stroke measures, but variation in care persists.^6^ Clinical decision support systems (CDSS) can improve adherence to recommended processes, although average absolute improvements across trials are modest and depend on design features such as workflow integration and patient-specific, actionable recommendations.^7–10^ In stroke, a multicenter trial of a CDSS in 77 hospitals improved care quality and reduced new vascular events at 3 months.^11^ Generative artificial intelligence assistants have had mixed effects on physicians’ diagnostic and management reasoning, and any decision aid carries the risk of automation bias, in which users follow erroneous advice.^7–9,12,13^ LUCID (Longitudinal Unified Clinical Intelligence for Decision support) encodes the 2026 AHA/ASA guideline as a directed acyclic graph of decision nodes spanning the acute stroke care pathway. For clinician-entered case features, it returns the applicable recommendations together with their COR, LOE, and supporting rationale. Whether decisions made with such a tool are guideline-concordant has not been studied, particularly for the recommendations with the highest stakes: Class 1 recommendations, for which the error is omission, and Class 3 recommendations, for which the error is commission. We conducted the LUCID Validation (LUCID-VAL) pilot vignette study to compare the guideline concordance of acute stroke decisions made with and without LUCID at the level of individual guideline decision points, and to assess decision confidence and workload.

## METHODS

### Study Design and Oversight

LUCID-VAL was a prospective pilot online clinical vignette study. The University of Pittsburgh Institutional Review Board approved the study (STUDY26030001), and participants provided electronic informed consent. The study is reported in accordance with, where applicable, the DECIDE-AI guideline for early-stage clinical evaluation of decision support systems.^14^ Data were collected between April and May 2026.

### Participants and Study Arms

Physicians involved in acute stroke care (vascular neurologists and vascular neurology fellows, neurointerventionalists, general neurologists and neurohospitalists, neurology residents in postgraduate year 2 or above, and emergency medicine physicians) were invited by email.

Participants were assigned by the study team to complete the vignettes alone (Arm A) or with access to LUCID (Arm B). Survey sessions were reconciled with participants’ self-generated identifiers; sessions with no vignette response, investigator test sessions, and empty or superseded duplicate sessions were removed **(Figure 1)**. Arm B participants received a brief orientation and could consult LUCID for any question. For each question, they entered their final answer as free text. Arm A participants answered without LUCID. A decision point was classified as made with LUCID when an Arm B participant reported consulting LUCID for that decision, and as made without LUCID when the participant was in Arm A or reported not using LUCID.

**Figure 1.**
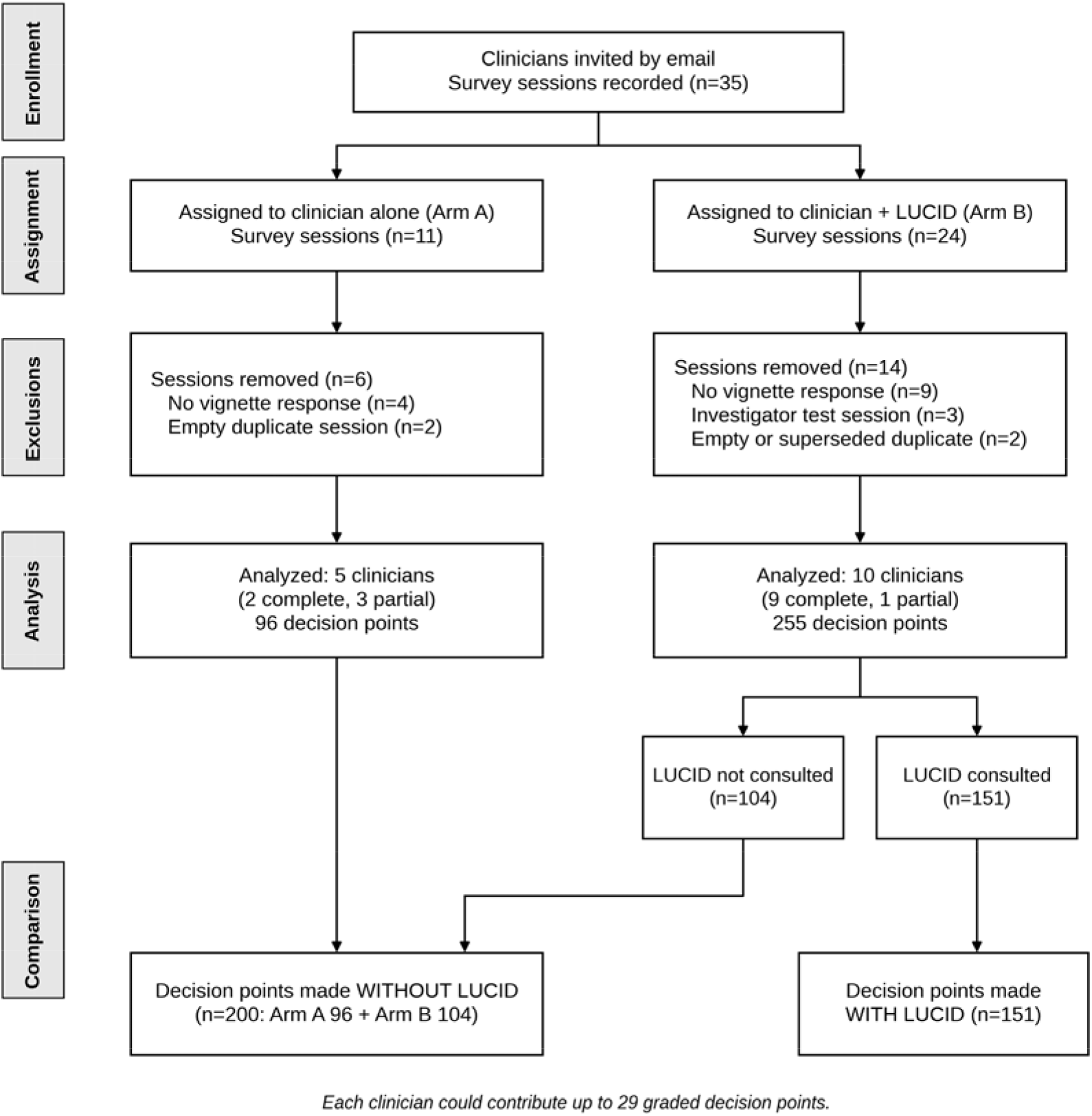
Study flow. Survey sessions recorded for each arm, sessions removed, and derivation of decision points made with and without LUCID.

### Clinical Vignettes and Decision Points

Eight vignettes spanned the acute stroke care pathway: prehospital care, emergency department evaluation, intravenous thrombolysis (IVT), endovascular thrombectomy (EVT), posterior and distal occlusions, post-reperfusion care, antithrombotic therapy, and inpatient management and complications. Each vignette posed 3 questions (24 questions); for each, participants provided a free-text answer, a rationale, and a confidence rating from 1 (very uncertain) to 5 (very confident). The vignettes and reference standard were developed by the investigators from the 2026 guideline.^4^ Each question was mapped to the guideline recommendation or recommendations that it tested and their COR. Because a single question could bundle recommendations of different classes, 4 questions were divided into 11 recommendation-specific nodes that were judged separately. The resulting 29 decision points **(Table S2)** comprised 10 Class 1, 5 Class 3: Harm, 6 Class 3: No Benefit, and 8 Class 2a, Class 2b, or split-class decision points.

### Outcomes

The primary outcome was full guideline concordance of each decision point. Secondary outcomes were at least partial concordance (score ≥0.5); full concordance within the Class 1, Class 3: Harm, and Class 3: No Benefit categories; omission errors (discordant responses to Class 1 decision points); commission errors (discordant responses to Class 3 decision points); decision confidence; and workload, measured with 4 NASA Task Load Index (NASA-TLX) subscales (mental demand, temporal demand, effort, and frustration; range 1–21).^15^ Decision confidence and workload were compared between study arms.

### Outcome Assessment

Each decision point was scored against the reference standard as fully concordant (1), concordant in its core decision but incomplete (0.5), or discordant or potentially harmful (0). For node-level decision points, each participant’s answer was graded separately at every node. Scoring was performed by the investigators, who were not blinded to study arm or LUCID use.

### Statistical Analysis

The unit of analysis was the decision point. Because decision points were clustered within clinicians, 95% CIs and 2-sided P values for risk differences (RDs) and mean differences were obtained by cluster bootstrap (4000 resamples of clinicians, stratified by arm).^16^ Omission and commission errors were compared with the Fisher exact test. Decision confidence was analyzed once per answered question, summarized as median (interquartile range [IQR]), and compared between Arm B and Arm A by cluster bootstrap of the mean difference. NASA-TLX scores, measured once per clinician at the end of the survey, were missing for 3 clinicians who stopped early (Arm A, 2; Arm B, 1). Missing scores were multiply imputed (100 imputations) by chained equations with predictive mean matching, using study arm, training status, and mean decision confidence as predictors, so that imputed values took observed scale values.^17,18^ Arm differences in mean scores were pooled by Rubin’s rules with small-sample degrees of freedom, and complete-case exact permutation tests were performed as a sensitivity analysis.^19^ Sensitivity analyses of the primary outcome analyzed undivided questions, restricted to clinicians who answered all questions, and omitted each clinician in turn. Exploratory analyses, considered descriptive, examined care nodes and clinician characteristics. P values for secondary outcomes were adjusted by the Holm method within families (recommendation classes, error types, and NASA-TLX subscales).

### Data Availability

Deidentified data and analysis code are available from the corresponding author on reasonable request.

## RESULTS

### Participants and LUCID Use

Of 35 recorded survey sessions, 20 were removed, leaving 15 clinicians: 5 in Arm A and 10 in Arm B **(Figure 1; Table S1)**. The clinicians contributed 351 decision points: 151 made with LUCID and 200 made without it (Arm A, 96; Arm B, 104). LUCID was thus consulted for 59.2% of Arm B decision points.

### Primary Outcome

Full guideline concordance was 99.3% (150 of 151) for decision points made with LUCID and 71.0% (142 of 200) for those made without it (RD, 28.3 [95% CI, 21.7–34.9] percentage points; P<0.001) **(Table 1)**. The only decision point made with LUCID that was not fully concordant received partial credit.

**Table 1.** Primary and Secondary Outcomes: Decision Points Made With vs Without LUCID and Clinician-Reported Outcomes by Study Arm.

| Outcome | With LUCID, n/N (%) | Without LUCID, n/N (%) | Effect (95% CI) | P | Holm P |
| --- | --- | --- | --- | --- | --- |
| **Primary outcome: fully concordant decision points** |  |  |  |  |  |
| All decision points | 150/151 (99.3) | 142/200 (71.0) | RD 28.3 (21.7 to 34.9) | <0.001 | — |
| **Partial-credit concordance** |  |  |  |  |  |
| At least partially concordant (score ≥0.5) | 151/151 (100.0) | 187/200 (93.5) | RD 6.5 (2.3 to 10.2) | 0.001 | — |
| **Concordance by Class of Recommendation** |  |  |  |  |  |
| Class 1 (10 decision points) | 45/45 (100.0) | 53/78 (67.9) | RD 32.1 (23.7 to 39.5) | <0.001 | <0.001 |
| Class 3: Harm (5 decision points) | 30/30 (100.0) | 23/29 (79.3) | RD 20.7 (5.0 to 34.8) | 0.018 | 0.018 |
| Class 3: No Benefit (6 decision points) | 36/36 (100.0) | 26/35 (74.3) | RD 25.7 (17.4 to 38.9) | <0.001 | <0.001 |
| **Errors (discordant response, score 0)** |  |  |  |  |  |
| Omission (Class 1) | 0/45 (0.0) | 2/78 (2.6) |  | 0.532 | 0.532 |
| Commission, Class 3: Harm | 0/30 (0.0) | 6/29 (20.7) |  | 0.011 | 0.032 |
| Commission, Class 3: No Benefit | 0/36 (0.0) | 4/35 (11.4) |  | 0.054 | 0.108 |
| Commission, any Class 3 | 0/66 (0.0) | 10/64 (15.6) |  | <0.001 | 0.002 |
| **Decision confidence (1–5) by study arm, one rating per question§: Arm B (clinician + LUCID) vs Arm A (clinician alone)** |  |  |  |  |  |
| Median [Q1–Q3] | 5 [4–5] (n=193) | 4 [3–5] (n=73) | Mean difference 0.5 (–0.2 to 1.5) | 0.242 | — |
| **NASA-TLX workload (1–21; lower is better) by study arm, mean after multiple imputation□: Arm B (n=10) vs Arm A (n=5)** |  |  |  |  |  |
| Mental demand | 11.8 | 14.7 | Difference –2.9 (–5.8 to –0.1) | 0.045 | 0.182 |
| Temporal demand | 7.5 | 14.0 | Difference –6.5 (–14.4 to 1.3) | 0.090 | 0.271 |
| Effort | 9.5 | 12.7 | Difference –3.2 (–8.5 to 2.2) | 0.205 | 0.409 |
| Frustration | 6.8 | 8.5 | Difference –1.7 (–8.8 to 5.4) | 0.601 | 0.601 |
| Composite (4 subscales) | 8.9 | 12.5 | Difference –3.6 (–7.8 to 0.6) | 0.084 | — |

Upper panels (decision points): with LUCID = Arm B decisions at which LUCID was consulted (n=151); without LUCID = Arm A decisions (n=96) and Arm B decisions made without consulting LUCID (n=104). RD indicates risk difference in percentage points; P values for differences are from cluster bootstrap (4000 resamples of clinicians). Error rows: Fisher exact test. Lower panels (study arms): the second and third columns show Arm B and Arm A, respectively. §One rating per answered question (Arm B, 193 ratings from 10 clinicians; Arm A, 73 ratings from 5 clinicians); mean difference and P value from cluster bootstrap. □Measured once per clinician; scores missing for 3 clinicians were multiply imputed (chained equations with predictive mean matching, 100 imputations); means and differences (Arm B – Arm A) pooled by Rubin's rules with small-sample degrees of freedom. Holm adjustment within families (recommendation classes; error types; NASA-TLX subscales).

### Secondary Outcomes

At least partial concordance was 100% (151 of 151) with LUCID versus 93.5% (187 of 200) without (RD, 6.5 [2.3–10.2] percentage points; P=0.001); without LUCID, 45 decision points were partially concordant and 13 were discordant. Full concordance was higher with LUCID in every recommendation class **(Table 1**; **Figure 2)**: Class 1, 100% versus 67.9% (RD, 32.1 [23.7–39.5]); Class 3: Harm, 100% versus 79.3% (RD, 20.7 [5.0–34.8]; P=0.018); and Class 3: No Benefit, 100% versus 74.3% (RD, 25.7 [17.4–38.9]); all remained significant after Holm adjustment. Without LUCID, the lowest concordance occurred for ambulance-based remote ischemic conditioning (1 of 5), graduated compression stockings (3 of 6), the blood pressure target after reperfusion (3 of 6), and thrombolytic agent and dose (3 of 6) **(Figure 3)**.

**Figure 2.**
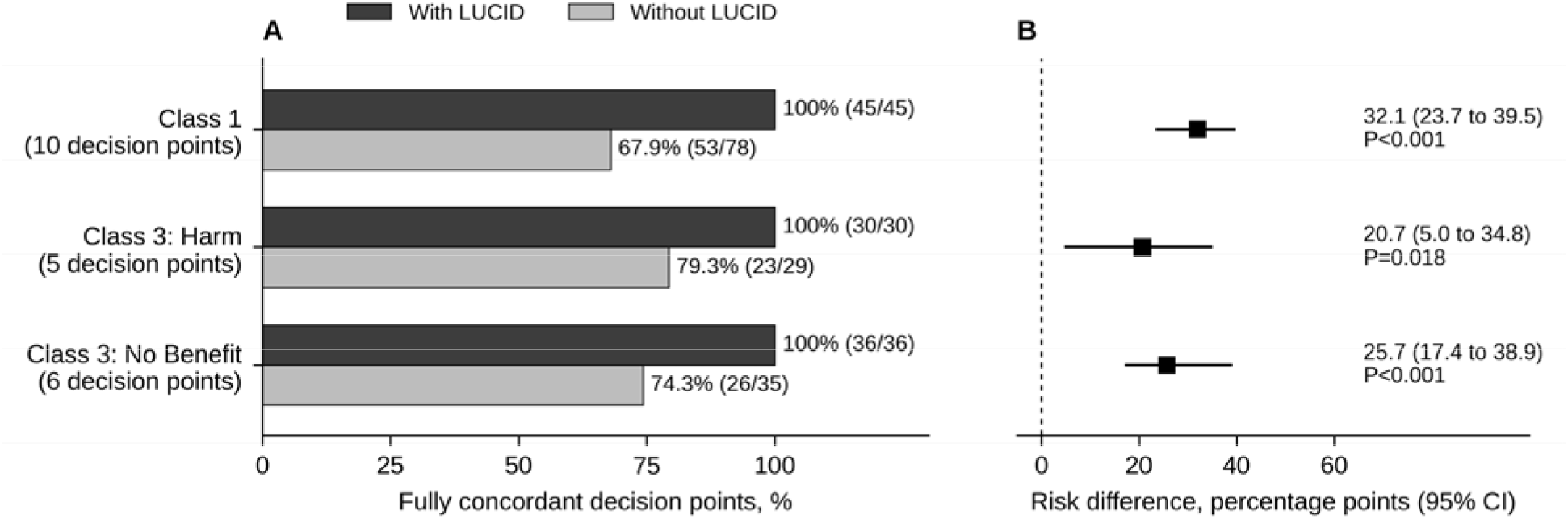
Guideline concordance with and without LUCID for Class 1 and Class 3 recommendations. A, Percentage of fully guideline-concordant decision points. B, Risk differences with 95% CIs and P values from cluster bootstrap resampling of clinicians. Without LUCID includes Arm A decisions and Arm B decisions made without consulting LUCID.

**Figure 3.**
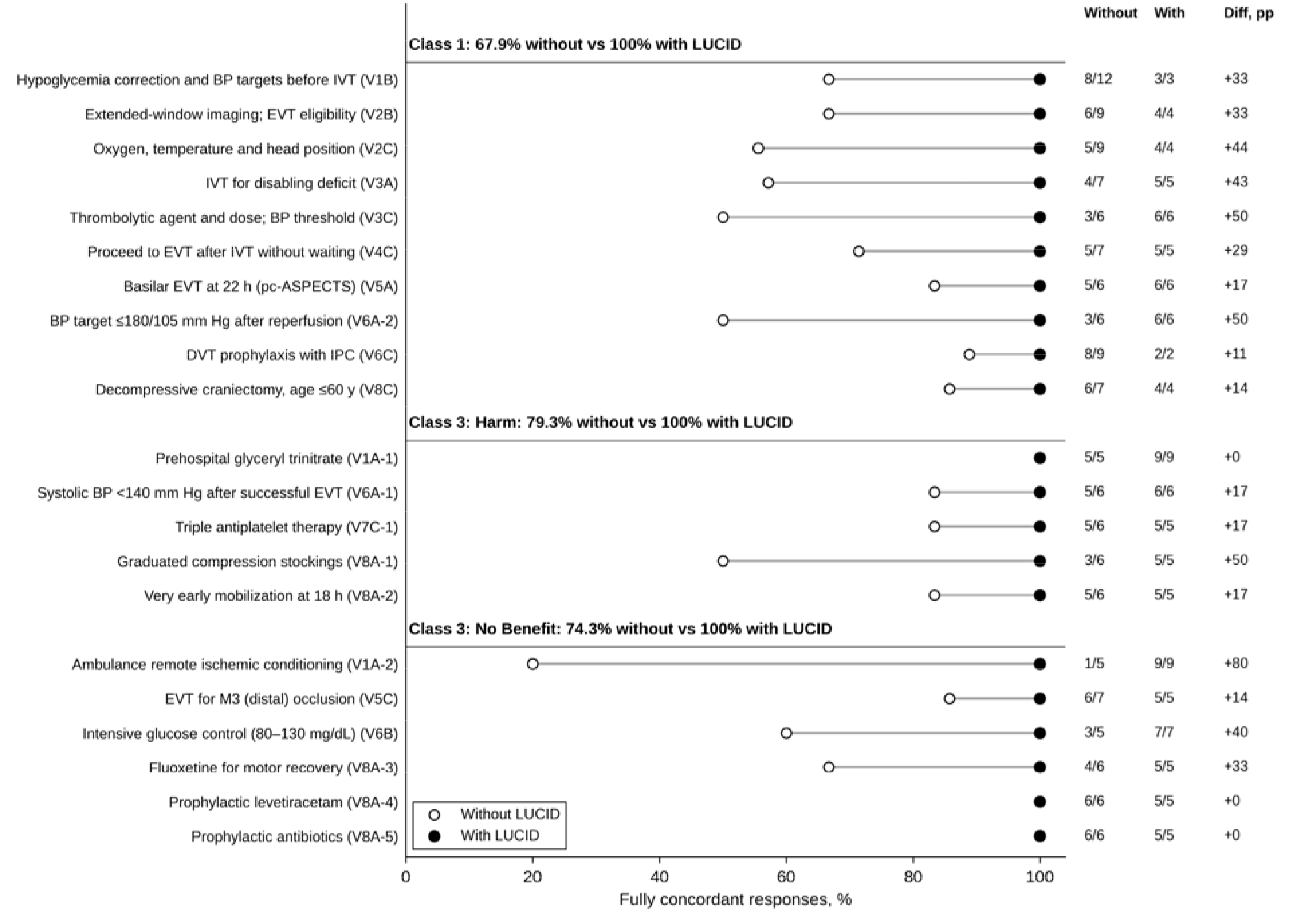
Concordance at each Class 1 and Class 3 decision point. Percentage of fully concordant responses without (open circles) and with (filled circles) LUCID for each of the 21 decision points testing a Class 1 or Class 3 recommendation, with numerators and denominators and the difference in percentage points. Per-point estimates are descriptive (2–12 responses per cell). BP indicates blood pressure; DVT, deep vein thrombosis; EVT, endovascular thrombectomy; IPC, intermittent pneumatic compression; and IVT, intravenous thrombolysis.

Commission errors occurred at 0 of 66 Class 3 decision points with LUCID and 10 of 64 (15.6%) without it (P<0.001; Holm-adjusted P=0.002), including 0 of 30 versus 6 of 29 for Class 3: Harm recommendations (P=0.011; Holm-adjusted P=0.032) and 0 of 36 versus 4 of 35 for Class 3: No Benefit recommendations (P=0.054). Omission errors were infrequent (0 of 45 versus 2 of 78; P=0.532).

Decision confidence was numerically higher in Arm B than in Arm A (median, 5 [interquartile range, 4–5] versus 4 [3–5]; mean difference, 0.5 [95% CI, −0.2 to 1.5]; P=0.24) **(Figure 4)**. After multiple imputation of missing NASA-TLX scores, workload was lower in Arm B for mental demand (mean, 11.8 versus 14.7; difference, −2.9 [95% CI, −5.8 to −0.1]; P=0.045) and borderline lower for temporal demand (7.5 versus 14.0; difference, −6.5 [−14.4 to 1.3]; P=0.090) and the composite score (8.9 versus 12.5; difference, −3.6 [−7.8 to 0.6]; P=0.084), whereas effort

**Figure 4.**
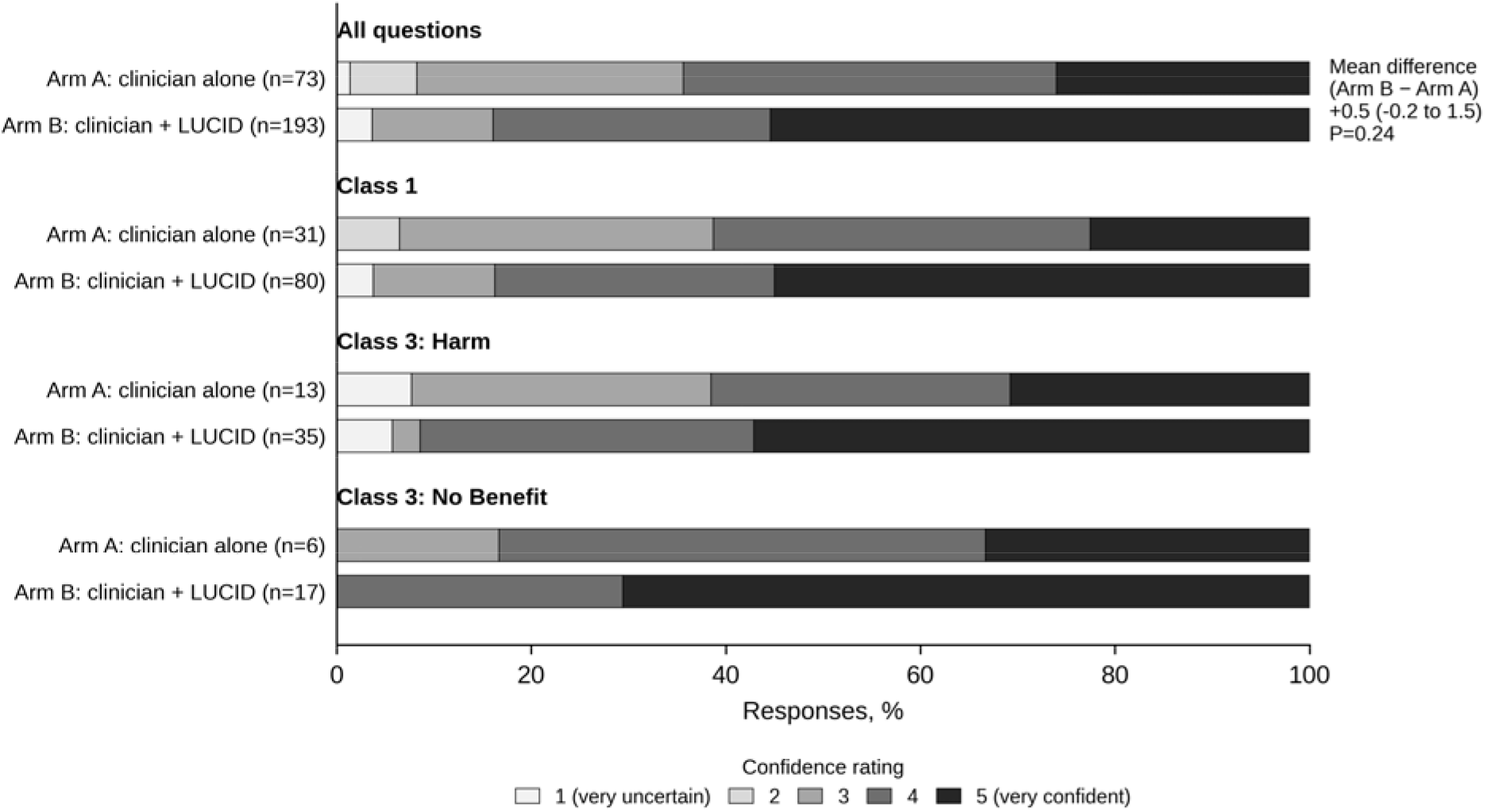
Decision confidence by study arm. Distribution of confidence ratings (1, very uncertain, to 5, very confident) for answered questions in Arm A (clinician alone) and Arm B (clinician + LUCID), overall and by Class of Recommendation. The mean difference (Arm B − Arm A), 95% CI, and P value are from cluster bootstrap resampling of clinicians.

(difference, −3.2 [−8.5 to 2.2]; P=0.20) and frustration (−1.7 [−8.8 to 5.4]; P=0.60) did not differ. No workload difference remained significant after Holm adjustment, and complete-case analyses showed the same direction for every subscale **(Table S5)**.

### Sensitivity and Exploratory Analyses

The primary result was consistent in the sensitivity analyses **(Table S3)**: the RD was 32.8 percentage points at the question level and 28.4 among clinicians who answered all questions, and it ranged from 26.2 to 30.6 when each clinician was omitted in turn. Concordance with LUCID exceeded that without it at all 8 care nodes and in every clinician subgroup that contained decisions made both with and without LUCID **(Table S4)**.

## DISCUSSION

In this pilot vignette study, 99.3% of decisions that clinicians made after consulting LUCID were fully concordant with the 2026 AHA/ASA guideline, compared with 71.0% of decisions made without it. The difference was present for every class of recommendation and at every care node. Without LUCID, clinicians endorsed interventions that the guideline designates as harmful or of no benefit in 10 of 64 instances; with LUCID, they did not do so once. Clinicians with access to LUCID also reported lower workload, most clearly for mental demand, although the workload differences did not withstand adjustment for multiple comparisons.

These findings matter because the recommendations with the greatest consequences for patients are concentrated in Class 1 and Class 3. The Class 3: Harm recommendations tested here rest on trials showing that prehospital glyceryl trinitrate, intensive systolic blood pressure lowering after successful thrombectomy, triple antiplatelet therapy, graduated compression stockings, and very early high-dose mobilization worsen outcomes or cause harm.^4^ Several of these recommendations run counter to intuitive practice, and unaided clinicians in this study endorsed several of them. A tool that presents the guideline class and level of evidence at the moment of decision may be particularly valuable for such counterintuitive or recently revised recommendations, including those on dual antiplatelet therapy beyond 24 hours, early anticoagulation in atrial fibrillation, and thrombectomy for medium- and distal-vessel occlusion.^4^ The errors observed without LUCID are consistent with the well-described gap between guideline publication and practice. Translation of evidence into routine care is slow and incomplete, and barriers include limited awareness of, or familiarity with, specific recommendations and the inertia of previous practice.^20–22^ These barriers are amplified when a guideline is long, frequently updated, and contains recommendations whose class has changed, as with several recommendations in the 2026 guideline. A decision aid that surfaces the applicable recommendation and its class at the moment of decision addresses awareness and recall directly, which may explain why the largest differences occurred for decisions on thrombolysis and thrombectomy, where recommendations have changed most rapidly.

Clinicians chose when to consult LUCID, so this comparison is observational, and decisions made with and without the tool may differ in ways other than tool use. The difference was nevertheless consistent across care nodes, clinician subgroups, and sensitivity analyses, including analyses restricted to clinicians who answered all questions and analyses omitting each clinician in turn. Residual confounding by indication, for example if clinicians consulted LUCID selectively for decisions they found more or less difficult, cannot be excluded.

Across controlled trials, CDSS improve the proportion of patients receiving desired care by approximately 6 percentage points on average. The larger difference observed here likely reflects the vignette setting, in which the tool was available at every decision, and LUCID’ provision of patient-specific, actionable recommendations, a feature associated with CDSS success. Our findings complement GOLDEN BRIDGE II, in which a stroke CDSS improved care quality and reduced vascular events, and contrast with trials of general-purpose large language models, which modestly improved management reasoning but did not improve diagnostic reasoning.^8–10,13,23–27^

A deterministic system that links each recommendation to its guideline source may also be easier to audit than generative models, whose outputs can be fluent but unsupported. Decision confidence was numerically higher in Arm B than in Arm A, but the difference was imprecise.

Physicians’ confidence is often poorly calibrated to their accuracy, particularly for difficult cases, and higher confidence is desirable only while the tool is correct: automation bias would lead clinicians to follow erroneous advice with the same confidence, so continued validation of the encoded logic against guideline updates is essential.^13,28^ Workload was lower among clinicians with access to LUCID, most clearly for mental demand, with a borderline reduction in temporal demand, which suggests that consulting the tool did not add burden. However, clinicians did not consult LUCID for 41% of decisions despite having access, and benefit accrues only when the tool is consulted; low uptake and override of decision support are common in practice, so faster data entry and integration with the electronic health record are priorities.^29^

This study has important limitations. First, it is a small pilot study of 15 clinicians. Second, clinicians chose when to consult LUCID, so the comparison of decisions made with and without LUCID is observational, and residual confounding cannot be excluded. Third, outcome assessment was not blinded and was performed by investigators with a financial interest in LUCID, and the 9 data-entry corrections confirmed by participants after the study applied only to decisions made with LUCID; with the initial scores, the difference in full concordance was 22.4 percentage points. Fourth, the reference standard was developed by the investigators, and several answer keys required interpretation. Fifth, LUCID use was self-reported, workload was measured once per clinician and was missing for 3 clinicians, and responses to vignettes may not reflect real-time clinical decisions, although vignettes are valid measures of the quality of physician practice.^30,31^ A larger confirmatory study should prespecify the decision-level comparison, use blinded, independent, duplicate outcome assessment applied identically to all responses, and evaluate LUCID integrated into clinical workflow.

In this pilot vignette study, decisions made with a guideline-encoded clinical decision support system were almost uniformly concordant with the 2026 AHA/ASA guideline, including recommendations against harmful interventions, whereas nearly 3 in 10 decisions made without it were not fully concordant. These findings require confirmation in a larger study with blinded outcome adjudication.

## Data Availability

Upon reasonable request from the authors.

## ARTICLE INFORMATION

## Acknowledgments

We thank the participating clinicians.

## Supplemental Material

Supplemental Methods; Tables S1–S5.

## Nonstandard Abbreviations and Acronyms

AHA/ASA: American Heart Association/American Stroke Association
CDSS: clinical decision support system
COR: Class of Recommendation
EVT: endovascular thrombectomy
IVT, intravenous: rombolysis
LOE: Level of Evidence
LUCID: Longitudinal Unified Clinical Intelligence for Decision support
LUCID-VAL: LUCID Validation study
NASA-TLX: NASA Task Load Index
RD: risk difference

